# Utility of Multimodal Longitudinal Imaging Data for Dynamic Prediction of Cardiovascular and Renal Disease: The CARDIA Study

**DOI:** 10.1101/2023.05.19.23290259

**Authors:** Hieu Nguyen, Henrique D. Vasconcellos, Kimberley Keck, John Carr, Lenore J Launer, Eliseo Guallar, João A.C. Lima, Bharath Ambale-Venkatesh

**Author notes:** Corresponding author: Bharath Ambale-Venkatesh, Assistant Professor of Radiology and Radiological Science MR 110, Radiology, Johns Hopkins University Baltimore, MD 21287.

## Abstract

**Background:** Medical examinations contain repeatedly measured data from multiple visits, including imaging variables collected from different modalities. However, the utility of such data for the prediction of time-to-event is unknown, and only a fraction of the data is typically used for risk prediction. We hypothesized that multimodal longitudinal imaging data could improve dynamic disease prognosis of cardiovascular and renal disease (CVRD).

**Methods:** In a multi-centered cohort of 5114 CARDIA participants, we included 166 longitudinal imaging variables from five imaging modalities: Echocardiography (Echo), Cardiac and Abdominal Computed Tomography (CT), Dual-Energy X-ray Absorptiometry (DEXA), Brain Magnetic Resonance Imaging (MRI) collected from young adulthood to mid-life over 30 years (1985-2016) to perform dynamic survival analysis of CVRD events using machine learning dynamic survival analysis (Dynamic-DeepHit, LTRCforest, and Extended Cox for Time-varying Covariates). Risk probabilities were continuously updated as new data were collected. Model performance was assessed using integrated AUC and C-index and compared to traditional risk factors.

**Results:** Longitudinal imaging data, even when being irregularly collected with high missing rates, improved CVRD dynamic prediction (0.03 in integrated AUC, up to 0.05 in C-index compared to traditional risk factors) from young adulthood followed up to midlife. Among imaging variables, Echo and CT variables contributed significantly to improved risk estimation. Echo measured in early adulthood predicted midlife CVRD risks almost as well as Echo measured 10-15 years later (0.01 C-index difference). The most recent CT exam provided the most accurate prediction for short-term risk estimation. Brain MRI markers provided additional information from cardiac Echo and CT variables that led to a slightly improved prediction.

**Conclusions:** Longitudinal multimodal imaging data readily collected from follow-up exams can improve CVRD dynamic prediction. Echocardiography measured early can provide a good long-term risk estimation, while CT/calcium scoring variables carry atherosclerotic signatures that benefit more immediate risk assessment starting in middle-age.

## Introduction

The rapidly expanding availability of large health data sets has fueled the growing research for more accurate risk prediction which holds much potential for preventive and monitoring strategies as well as improved disease understanding. In many scenarios, imaging data are collected over various modalities (multimodal) such as Echocardiography, Magnetic Resonance Imaging, and Computed Tomography, and repeatedly measured in multiple follow-up exams. Multimodal longitudinal imaging data could provide a more comprehensive description of the body and the development of organ functions and structures over time. In cardiology, numerous imaging markers for subclinical atherosclerosis have been demonstrated to be independently predictive of cardiovascular events^1,2,3^ and cardiac dysfunction.^4,5^ Many published works have focused on a few imaging variables that are low-dimensional, single-modal,^2,4– 6^ or cross-sectional.^7^ The utility of high-dimensional, multimodal, and longitudinal imaging data has not been investigated.

Cox Proportional Hazards (Cox-PH) is among the most popular methods for survival analysis but Cox-PH is not suitable for high-dimensional data with repeated measures. The extended version of Cox-PH that can work with time-varying covariates is still limited because of the high number of variables, nonlinearity of variables, and requirement of data with no missingness.^8^ Machine learning (ML) approaches such as Random Survival Forest^9^ can mitigate some of Cox’s limitations, but many ML methods are limited to static prediction and cannot perform dynamic survival analysis. In static prediction, the model does not automatically update as new observations are collected, (new data would require refitting an existing model or training a new model). Unlike static prediction, a dynamic survival analysis model automatically updates predicted survival probabilities as additional longitudinal observations are collected, and the model is trained only once. The ability to dynamically update risk as new information rolls in makes dynamic survival analysis attractive.^10,11^

In this work, we demonstrated the utility of dynamic prediction of Cardiovascular and Renal Disease (CVRD) using high-dimensional, multimodal, longitudinal imaging. Data were collected in CARDIA, which is a large epidemiological study of Black and White young adults followed up over 30 years. We also identified the most important imaging predictors for CVRD in the CARDIA cohort.

## Methods

### Study Population and Outcome

The design of the CARDIA study (Coronary Artery Risk Development in Young Adults) has been described elsewhere.^12^ Briefly, CARDIA is a prospective, observational cohort study of 5114 (originally 5115, one person withdrew consent) White and Black men and women aged 18 to 30 years, at four centers in the United States. The cohort is approximately balanced regarding age, race, sex, and educational level. Participants have been followed since 1985, with regular exam visits scheduled every 2-5 years. Each exam has collected a wide variety of variables believed to be related to heart disease. The institutional review board of each participating institution approved the study protocol and all participants gave informed consent.

The outcome of this study is cardiovascular and renal disease (CVRD), and the first CVRD event was used as the endpoint. These events were adjudicated through August 2019. The primary composite outcome was incident cardiovascular disease and renal disease, which included coronary heart disease (CHD, myocardial infarction, acute coronary syndrome, or CHD death, including fatal myocardial infarction), stroke, transient ischemic attack, hospitalization for heart failure, intervention for peripheral arterial disease, end-stage renal disease, or death from cardiovascular or renal causes. Participants who died from a non-CVRD cause were censored at the time of death in the survival models.

### Imaging Markers

CARDIA follow-up exams collect various imaging variables from different sources, such as Echocardiography (Echo), Computed Tomography (CT), Carotid Ultrasonography (CARTD), Dual-Energy X-ray Absorptiometry (DEXA), and Brain Magnetic Resonance Imaging (MRI). The extracted imaging variables have a high degree of sparsity and irregularity, reflecting real-world data. Echo was performed as part of the core study in Y5, Y25, and Y30 and as a substudy in Y10; CT was conducted in Y10 as a substudy and in Y15, Y20, and Y25 as the core study, and brain MRI was acquired in Y25 and Y30 as a substudy. Figure 1 shows an overview of imaging markers used in this study and Table S1 shows a detailed list of when the measures were collected. Data collection protocols for each imaging modality are available on the CARDIA study website.^13^ We used the longitudinal imaging CARDIA data from all exam years to develop prediction models.

**Figure 1.**
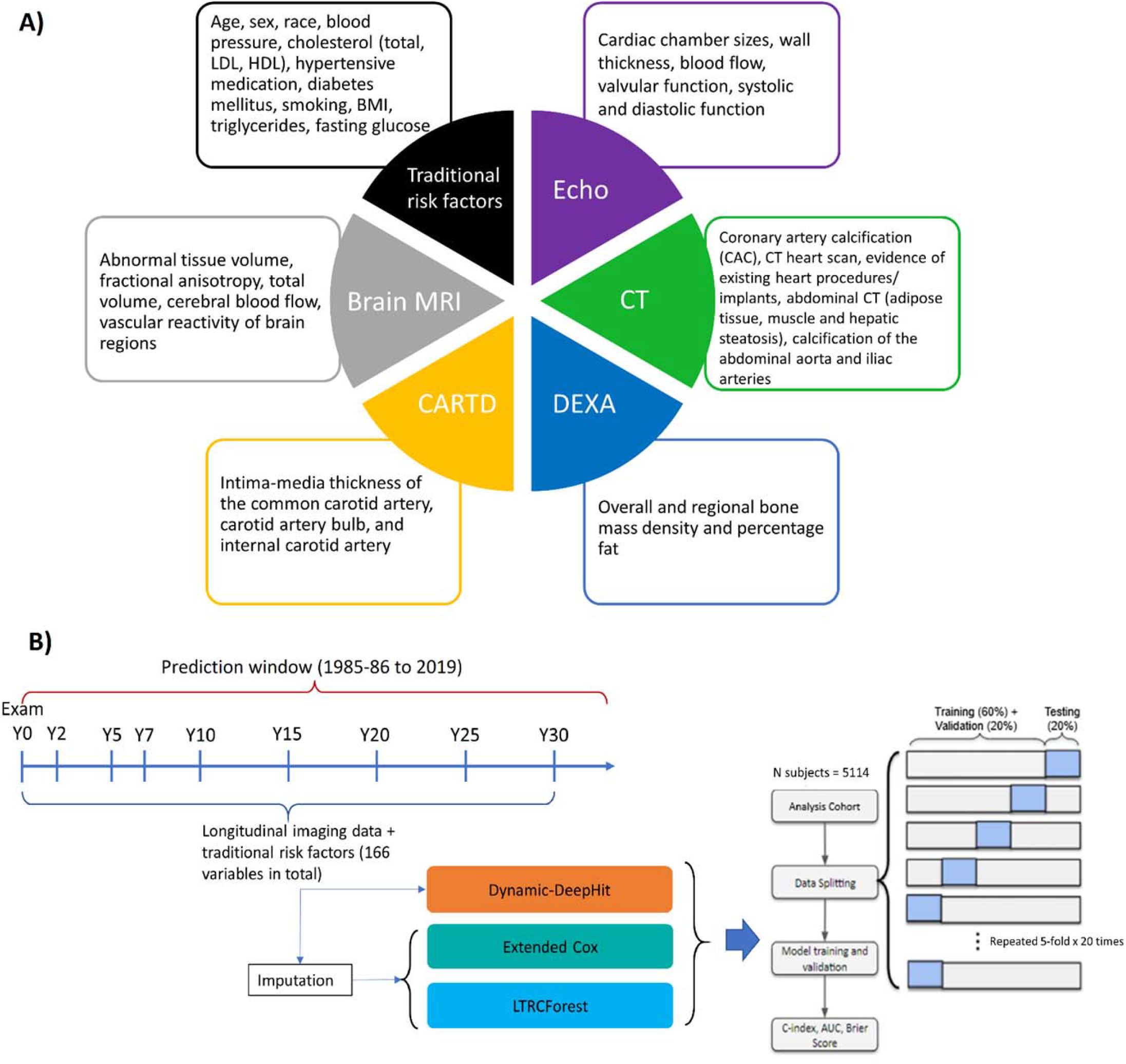
A) Overview of the multimodal data used for prediction. B) Methodology framework visualization for dynamic survival analysis. Echo: echocardiography, CT: Computed Tomography, CARTD: Carotid Artery Ultrasonography, DEXA: Dual-Energy X-ray Absorptiometry, MRI: Magnetic Resonance Imaging.

Variables were pre-filtered with help from domain experts (clinicians who performed image reading daily). Other exclusion criteria include removing duplicated variables across modalities, variables available in too few subjects and variables with poor documentation. Overall, we included a total of 151 longitudinal imaging markers. In addition, we also included 15 traditional risk factors: nine variables from the AHA/ACC ASCVD risk scores and six additional risk factors (diastolic blood pressure - DBP, body-mass index - BMI, taking cholesterol-lowering medications, low-density lipoproteins - LDL, triglycerides, and fasting glucose).

### Statistical Analysis

#### Model Training and Evaluation

Figure 1 shows the schematic of the statistical analysis procedures. All the models were trained and evaluated on the same cohort by 5-fold x 20 times cross-validation scheme. For each time the whole data was split, 20% of the data was used for testing, and the remaining 80% was further divided into training and validation sets. The training sets were used to fit the models, the validation sets were for hyperparameter tuning, and the testing sets were for assessing model performance.

Stratified sampling by event was conducted to ensure the same ratio of events to non-events across the splits.

#### Modeling Methods

We used three algorithms to model data for dynamic survival analysis. To issue dynamically updated survival predictions the data requires methods that can incorporate high-dimensional, longitudinal data comprising various repeated measurements with varying degrees of missingness. The main algorithm we used is Dynamic-DeepHit.^11^ Dynamic-DeepHit is a deep learning-based approach that issues dynamically updated survival predictions without making any assumptions about the underlying processes. Briefly, Dynamic-DeepHit consists of two subnetworks: a shared subnetwork with a recurrent neural network architecture that handles longitudinal measurements and predicts the next measurements of time-varying covariates, and a second subnetwork with cause-specific survival networks of fully connected layers that relates the longitudinal data to the survival outcome. Dynamic-DeepHit trains by minimizing the loss function which comprises three losses: a survival loss of log-likelihood of joint time-to-event distribution, a ranking loss that adapts the idea of concordance that encourages correct ordering of participants based on their time-to-event, and a step-ahead prediction loss that encourages correct prediction of longitudinal covariates for the next time step. Detailed description of Dynamic-DeepHit can be found elsewhere.^11^

We employed two additional methods for dynamic prediction, namely Left-Truncated-Right-Censored Forest (LTRCforest)^14^ and Extended Cox for Time-dependent Covariates.^8^ Briefly, the Extended Cox is an extension of the fully parametric Cox-PH that assumes the variable values remain constant from the last observed value until updated. To handle a large number of input covariates, LASSO penalization^15^ was employed. LTRCforest is an extension of the non-parametric ML method conditional forest (Cforest) for time-varying covariates. Since LTRCforest and Extended Cox require fully available data, missing data were imputed before being input into these models using Multiple Imputation by Chained Equations (MICE) for multilevel data.^16,17^

As a benchmark, we also fit static survival models at three time points (5 years, 15 years, and 25 years after baseline) to compare with the dynamic survival models.^18^ The idea is similar to landmarking approaches, in which a survival model is fit to the subjects who are still at risk at the landmarking time. For consistency in comparison, we used Dynamic-DeepHit and made an artificial cut-off at the landmarking time (meaning, covariate measurements after Y5, Y15, and Y25 were excluded from the static model at landmarking time 5 years, 15 years, and 25 years after baseline, respectively). Only measurements collected before the landmarking time were included.

#### Importance of imaging subsets and variables

To evaluate the effect of each imaging variable subset on CVRD prediction, we built five Dynamic-DeepHit models, each with traditional risk factors and imaging markers from a single modality. We also built a model with all imaging markers from all modalities and a reference model with only traditional risk factors. Additionally, to assess the complementary effects of multiple imaging subsets, we built 25 additional models representing all possible combinations of five imaging subsets. In total, 32 models were built (Table S2).

To examine the effect of imaging variables collected at different ages, we also built separate models for each imaging subset that included variables collected from each exam and excluded measurements collected outside of the exam.

Additionally, the importance of each imaging marker was quantified using permutation importance, similar to permutation testing,^19^ for the model trained on all variables. The longitudinal trajectories were permuted among participants, and the drop in the C-index of permuted variables to the C-index of the original dataset was used as the ranking criteria for variable importance. A bigger drop in C-index indicated a more important variable, and a minimal drop suggested that the variable was not important, as changing the variable value did not change model performance. Variables with the same difference in C-index were assigned the same ranking.

#### Performance Evaluation

Model performance was quantified using the time-dependent area under the receiver-operating curve (AUC) accounting for censorship^20^ and the time-dependent concordance index that accounted for censoring distribution.^21^ In addition, the integrated AUC (iAUC) was used to quantify all time-varying AUCs as one number.^22^ Statistical significance was evaluated using Wilcoxon rank sum test.

## Results

A total of 5114 participants were included in the analysis. Table 1 describes the characteristics and number of remaining participants in the cohort over nine follow-up exams. The mean age was 25 years old in CARDIA Y0 Exam (baseline) and 55 years old in the last exam (Y30). The cohort consisted of 46% male, 52% black, and 48% white. Over 30 years of follow-up, 3358 came back for Y30, the averaged SBP and DBP (systolic and diastolic blood pressure) increased and so was the use of hypertensive medication. BMI, total cholesterol, high-density lipoprotein (HDL), and use of cholesterol-lowering medication increased. The prevalence of diabetes also increased, while the number of smokers decreased. By the end of follow-up, 375 participants (7.3%) had developed CVRD. The cumulative incidence of CVRD is shown in Figure S1, with very few events happening before Y10 Exam, while the event rate curve is almost exponential after Y20 Exam.

**Table 1.** Characteristics of the study population over time.

|  | **Exam**<br>**Y0** | **Exam**<br>**Y2** | **Exam**<br>**Y5** | **Exam**<br>**Y7** | **Exam**<br>**Y10** | **Exam**<br>**Y15** | **Exam**<br>**Y20** | **Exam**<br>**Y25** | **Exam**<br>**Y30** |
| --- | --- | --- | --- | --- | --- | --- | --- | --- | --- |
| **No. subjects**<br>**examined** | (N=5114<br>) | (N=4624<br>) | (N=4352<br>) | (N=4085<br>) | (N=3948<br>) | (N=3671<br>) | (N=3549<br>) | (N=3499<br>) | (N=3358<br>) |
| **Age** | 24.8<br>(3.63) | 26.8<br>(3.60) | 29.9<br>(3.60) | 31.9<br>(3.58) | 34.9<br>(3.61) | 40.0<br>(3.60) | 45.0<br>(3.59) | 50.0<br>(3.59) | 55.0<br>(3.55) |
| **Sex (%Male)** | 2327<br>(46%) | 2089<br>(45%) | 1958<br>(45%) | 1836<br>(45%) | 1755<br>(44%) | 1619<br>(44%) | 1527<br>(43%) | 1505<br>(43%) | 1352<br>(43%) |
| **Race** |  |  |  |  |  |  |  |  |  |
| **Black** | 2651<br>(52%) | 2300<br>(50%) | 2132<br>(49%) | 1987<br>(49%) | 1938<br>(49%) | 1741<br>(47%) | 1649<br>(47%) | 1632<br>(47%) | 1510<br>(48%) |
| **White** | 2463<br>(48%) | 2323<br>(50%) | 2219<br>(51%) | 2098<br>(51%) | 2010<br>(51%) | 1930<br>(53%) | 1884<br>(53%) | 1842<br>(53%) | 1639<br>(52%) |
| **Systolic blood**<br>**pressure (SBP)** | 110<br>(10.9) | 108<br>(10.8) | 108<br>(11.6) | 109<br>(12.4) | 110<br>(12.8) | 113<br>(14.9) | 117<br>(15.2) | 120<br>(16.2) | 121<br>(16.6) |
| **Diastolic blood**<br>**pressure (DBP)** | 68.6<br>(9.62) | 67.4<br>(9.67) | 69.2<br>(10.2) | 69.3<br>(10.3) | 72.4<br>(10.2) | 74.5<br>(11.6) | 73.1<br>(11.5) | 74.9<br>(11.2) | 74.0<br>(11.1) |
| **Use of hypertensive**<br>**medication** | 115<br>(2%) | 41 (1%) | 70 (2%) | 81 (2%) | 135<br>(3%) | 292<br>(8%) | 615<br>(17%) | 936<br>(27%) | 1033<br>(33%) |
| **Body Mass Index**<br>**(BMI)** | 24.5<br>(5.05) | 25.2<br>(5.38) | 26.2<br>(5.91) | 26.8<br>(6.13) | 27.5<br>(6.54) | 28.8<br>(6.84) | 29.5<br>(7.25) | 30.2<br>(7.19) | 30.5<br>(7.19) |
| **Total cholesterol**<br>**(mg/dL)** | 177<br>(33.5) | 183<br>(35.2) | 178<br>(34.3) | 177<br>(34.3) | 178<br>(34.6) | 185<br>(35.8) | 186<br>(34.9) | 192<br>(36.9) | 191<br>(38.1) |
| **Use of cholesterol-**<br>**lowering medication** | 0 (0%) | 0 (0%) | 11 (0%) | 10 (0%) | 19 (0%) | 88 (2%) | 311<br>(9%) | 540<br>(16%) | 632<br>(20%) |
| **Total HDL**<br>**cholesterol (mg/dl)** | 53.2<br>(13.2) | 54.8<br>(14.1) | 53.3<br>(14.2) | 52.1<br>(14.2) | 50.3<br>(14.0) | 50.7<br>(14.6) | 54.2<br>(16.7) | 58.0<br>(18.0) | 59.9<br>(19.0) |
| **Total LDL**<br>**cholesterol (mg/dl)** | 109<br>(31.3) | 113<br>(33.3) | 108<br>(32.1) | 108<br>(31.6) | 109<br>(32.1) | 113<br>(32.3) | 110<br>(32.0) | 112<br>(32.8) | 110<br>(33.2) |
| **Triglycerides**<br>**(mg/dl)** | 72.9<br>(48.5) | 78.9<br>(53.4) | 80.8<br>(72.2) | 86.4<br>(75.7) | 92.1<br>(74.7) | 105<br>(92.8) | 109<br>(79.9) | 114<br>(85.9) | 108<br>(99.5) |
| **Fasting glucose**<br>**(mg/100 ml)** | 82.6<br>(16.3) | NA<br>(NA) | NA<br>(NA) | 90.1<br>(19.4) | 88.2<br>(20.4) | 86.7<br>(21.0) | 98.0<br>(26.4) | 99.5<br>(28.7) | 102<br>(32.0) |
| **Diabetes Mellitus** | 43 (1%) | 55 (1%) | 83 (2%) | 142<br>(3%) | 173<br>(4%) | 212<br>(6%) | 273<br>(8%) | 373<br>(11%) | 442<br>(14%) |
| **Smoking now** | 1546<br>(30%) | 1359<br>(29%) | 1243<br>(29%) | 1096<br>(27%) | 1005<br>(25%) | 808<br>(22%) | 677<br>(19%) | 588<br>(17%) | 436<br>(14%) |
| **CVRD event** | 0 (0%) | 0 (0%) | 2 (0%) | 7 (0.2%) | 14<br>(0.4%) | 56<br>(1.5%) | 112<br>(3%) | 208<br>(6%) | 313<br>(10%) |

### Dynamic versus static prediction

Figure 2 shows the performance over time for dynamic prediction versus static prediction. The dynamic survival model using Dynamic-DeepHit trained on all 166 variables had a C-index of 0.80-0.82 before Y20 and slightly dropped to 0.78 by the last time point, 33 years after baseline. The C-index of the dynamic survival model is higher than that of the static survival models across all time points, by a margin of 0.01-0.06. Unlike the static survival models that required a separate model at each landmarking time, the dynamic model was only trained once and automatically updated survival probabilities as a new measurement updated from a follow-up exam.

**Figure 2.**
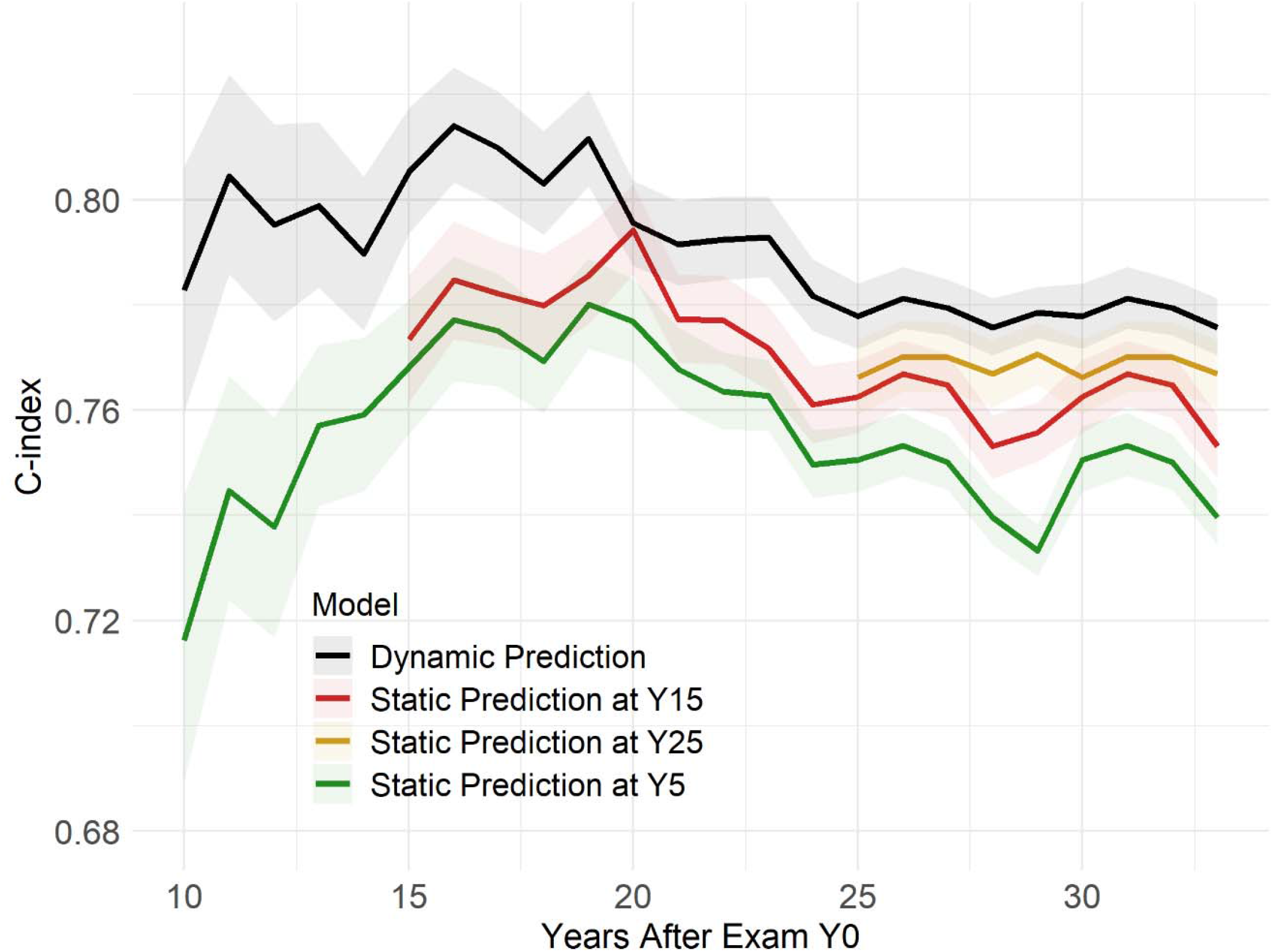
Dynamic vs. static prediction.

### Comparison of modeling methods

Figure S2 shows the performance of dynamic models trained with different algorithms (Dynamic-DeepHit, LTRCforest, Extended Cox, and Extended Cox penalized by LASSO). For both cases (trained on all variables and trained on traditional risk factors), Dynamic-DeepHit trained on unimputed data and LTRCforest trained on imputed data are consistently the best, whereas Extended Cox consistently underperformed (∼0.05-0.10 lower in C-index) for the model with all variables and 3-5% lower for model with only traditional risk factors).

### Predictive gain of modalities

Figure 3 shows the predictive gain from each imaging subset over time and on average over 25 cross-validation folds. The best model was the one that included all imaging subsets, while the worst-performing was the one using only traditional risk factors (baseline) and using traditional risk factors plus CARTD variables (0.74 C-index at end of follow-up). Using all imaging markers resulted in up to a 5% increase in C-index and 3% in iAUC. The model utilizing only CT variables was only slightly below (1%) the model using all imaging variables, which helped elevate performance since Y10 Exam with a more apparent gain after Y20 Exam when more CT variables were collected. The model trained on Echo variables shows that the inclusion of Echo variables in addition to traditional risk factors helped increase prediction accuracy throughout the entire follow-up period by ∼1.5-2% in C-index. DEXA variables improved performance very slightly up to Y25 by C-index (<0.01 absolute difference) and had negligible gain in terms of iAUC. Brain MRI variables, collected at Y25 and Y30 Exam, helped boost CRVD prediction performance by 0.01-0.02 C-index gain.

**Figure 3.**
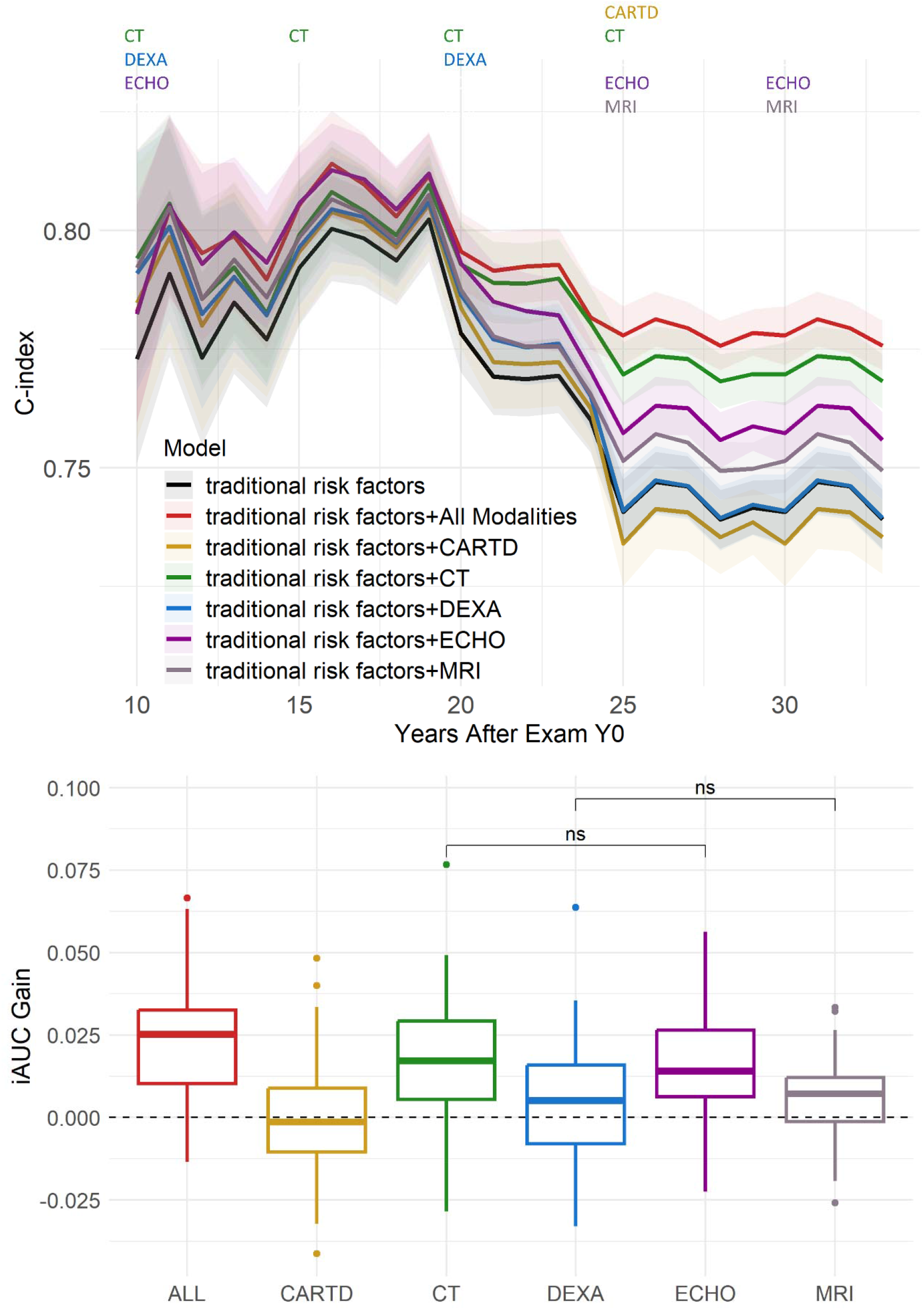
Predictive Gain from Imaging Variables of Different Modalities. Top: performance over time. The colored texts at the top indicate which imaging modalities were collected at each exam. Bottom: integrated AUC (iAUC) Gain with respect to the baseline model trained on only traditional risk factors. All pairwise hypothesis tests are significant (including All vs. ECHO and All vs. CT), unless otherwise denoted with ‘ns’ (‘non-significant’).

Table S2 shows the averaged iAUC gain from each of the 32 exhaustive combinations of 5 imaging variable subsets with respect to the baseline model. Aside from the best model using all imaging markers, the second-best subset is a combination of Echo, CT, and Brain MRI markers with an averaged iAUC gain of 0.027. A combination of Echo and CT variables resulted in a 0.022 iAUC gain, and the largest gain from a single imaging subset was from CT variables (0.014 iAUC gain).

### Temporal Importance

Figure 4 quantifies the importance of early versus late measurements on CRVD prediction in two imaging subsets with the most influence on prediction: Echo and CT. For Echo, the model trained on only early Echo measurements (collected in Y5 Exam as a core study and partially in Y10 Exam as a substudy) had just as good overall performance (iAUC = 0.78) as the model trained on Echo variables collected later in life (Y20 and Y25 Core Exams) (iAUC=0.78). For longer-term risk prediction (25 years after Y0), the C-index of the model trained on early Echo was only 1% less than that of the model trained on late Echo. Regarding CT variables, which were collected in Y10 as a substudy and in Y15, Y20, and Y25 as core studies, the most recent CT exam provided the most accurate prediction, as evidenced by the immediate bump in the C-index after each CT Exam. The most prominent bump is right after CT Y25 which resulted in a 3% increase in C-index compared to using CT variables from Y20 or earlier (p<0.001).

**Figure 4.**
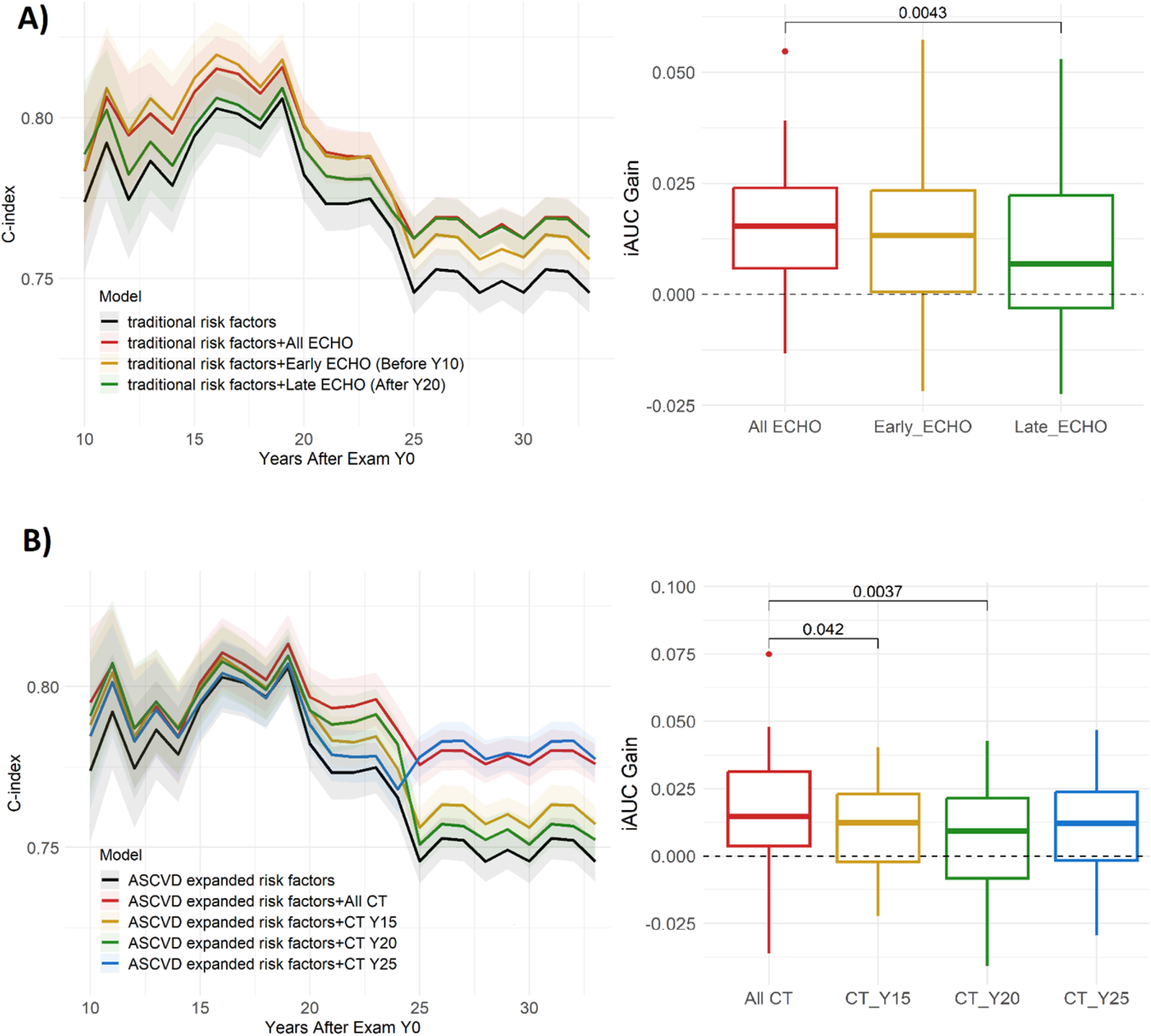
Effects of early vs. late imaging measurement. A) Early vs. Late Echo, B) Early vs. Late CT. Early Echo provided good overall gain for long-term risk estimation, compared to Late Echo. For CT, the most recent CT exam provides the most accurate prediction, evident by immediate bumps after each CT Exam, especially CT Y25. P-values of significant pairwise hypothesis tests are shown.

### Variable importance

In addition to quantifying the importance of variable subsets relative to each other, variable-level importance was quantified. Table 2 shows the top 20 ranked variables at three representative times: 15 years, 25 years, and 33 years (endpoint) after Y0. Total cholesterol and low-density lipoprotein cholesterol (LDL) were consistently the most important predictors of CVRD. Most of the top 20 variables were either collected by Echo and CT, attesting to their importance to CVRD prediction and consistent with the results in Figure 2. Chronological age is an important variable for year 15 (mean age=40) but not among the top 20 as participants got older. The top Echo and CT variables have similar relative variable importance in the rankings.

**Table 2.** Variable Importance Ranking (top 20)

| Year 15 |  |  |  | Year 25 |  |  |  | Year 33 |  |  |  |
| --- | --- | --- | --- | --- | --- | --- | --- | --- | --- | --- | --- |
| Rank | RVI | Modality | Variable | Rank | RVI | Modality | Variable | Rank | RVI | Modality | Variable |
| 1 | 1.00 | Traditional | Total Cholesterol | 1 | 1.00 | Traditional | Total Cholesterol | 1 | 1.00 | Traditional | Total Cholesterol |
| 2 | 0.94 | Traditional | LDL | 2 | 0.94 | Traditional | LDL | 2 | 0.91 | Traditional | LDL |
| 3 | 0.63 | Traditional | Age | 3 | 0.55 | Echo | Circumferential Peak Strain (%) | 3 | 0.74 | Echo | Mitral Regurgitation (Moderate Or Severe) |
| 4 | 0.59 | Echo | Vent Septal Thickness - Systole | 4 | 0.55 | Echo | LV Internal Dim In Diastole | 4 | 0.64 | Echo | Tricuspid Regurgitation (RVsp > 40 Mmhg) |
| 5 | 0.59 | Echo | Vent Septal Thickness - Diastole | 5 | 0.54 | Echo | LV Internal Dim In Systole | 5 | 0.63 | Echo | Mitral Valve E-wave To A-wave Ratio |
| 5 | 0.59 | Echo | 2D: LV Volume In Systole | 6 | 0.51 | Echo | Tricuspid Annular Peak Systolic Excursion | 6 | 0.63 | Echo | Aortic Root Dimension |
| 6 | 0.58 | Traditional | Smoking status | 7 | 0.51 | Echo | Aortic Root Dimension | 7 | 0.62 | Echo | Global LV Longitudinal Peak Strain (%) |
| 6 | 0.58 | CT | Agatston Score Left Anterior Descending | 8 | 0.50 | Echo | Vent Septal Thickness - Systole | 8 | 0.61 | Echo | LV Ejection Fraction |
| 7 | 0.58 | Echo | LV Internal Dim In Systole | 9 | 0.49 | Echo | LV Volume In Systole | 9 | 0.61 | MRI | Volume Of Total Brain |
| 8 | 0.58 | CT | Volume Of Lesions LAD | 10 | 0.48 | CT | Standard Deviation Attenuation Of Liver | 10 | 0.61 | Echo | LV Stroke Volume |
| 9 | 0.58 | Echo | Aortic Root Dimension | 11 | 0.48 | Echo | Vent Septal Thickness - Diastole | 11 | 0.60 | Echo | LV Internal Dim In Systole |
| 10 | 0.58 | Echo | AFVI/EFVI Ratio | 12 | 0.48 | Echo | LV Post Wall Thickness - Diastole | 12 | 0.60 | Echo | Circumferential Peak Strain (%) |
| 10 | 0.58 | CT | No. Of Lesions Left Anterior Descending | 13 | 0.47 | Echo | LV Internal Dimension Systole | 13 | 0.60 | Echo | LV Volume In Systole |
| 11 | 0.58 | DEXA | Hip: Total BMD | 14 | 0.47 | CT | Calcium Mass Of Lesions Infrarenal Abd. Aorta | 14 | 0.59 | MRI | Abnormal Tissue Volume In Gray Matter |
| 12 | 0.58 | CT | No. Of Lesions All Coronary | 15 | 0.47 | CT | Volume Of Lesions Right Coronary | 15 | 0.59 | Echo | Vent Septal Thickness - Diastole |
| 12 | 0.58 | Echo | LV Ejection Fraction | 16 | 0.47 | MRI | Volume Of Total Brain | 16 | 0.59 | CT | Stent Present |
| 13 | 0.58 | CT | Volume Of Lesions All Coronary | 16 | 0.47 | CT | Valve Replacement Present |  | 0.59 | CT | Calcium Mass Of Lesions LAD |
| 14 | 0.58 | CT | Mean Of Lesions Of Whole Heart | 17 | 0.46 | CT | No. Of Lesions Right Common Iliac | 18 | 0.59 | CT | Coronary Artery Bypass Graft |
| 14 | 0.58 | CT | Calcium Mass Of Lesions LAD | 18 | 0.46 | Echo | Doppler Tissue Doppler RV S-wave Velocity | 19 | 0.59 | CT | Cardiac Surgery Present |
| 14 | 0.58 | CT | Mean Of Lesions Left Circumflex | 19 | 0.46 | MRI | Abnormal Tissue Volume In Gray Matter | 20 | 0.58 | MRI | CSF Volume |

RVI: relative variable importance; LDL: low-density lipoprotein; LV: left ventricular; RV: right ventricular; AFVI/EFVI: atrial to early diastolic flow velocity integral; BMD: bone mass density; LAD: left anterior descending artery; CSF: cerebrospinal fluid.

## Discussion

In this work, we investigated the utility of high-dimensional longitudinal imaging data of five modalities, separately and together, for dynamic prediction of CVRD in young adults in a multi-centered cohort followed up over 30 years. We used the entirety of imaging variables over all exam years for continuously updated predictions of risk. The results suggest that longitudinal imaging data, even when irregularly collected and having high missing rates, improved CVRD dynamic prediction (3% iAUC, up to 5% C-index in midlife). Among different subsets of imaging markers, Echo and CT contributed to most of the improved risk estimation. Brain MRI variables contributed additional information that slightly improved prediction when they were collected. DEXA and Carotid IMT contributed little to none to CVRD prediction, even though they could be helpful in other aspects of clinical and epidemiological research. In terms of the effects of imaging markers measured early or late in life, the results suggested that Echo measured in early adulthood could predict long-term CVRD risks almost as well as Echo measured 10-15 years later. For CT, the most recent CT exam provides the most accurate prediction for short-term CVRD risk estimation. The results also suggest that the prediction ability of models decreased over time, particularly so between the ages of 40 and 50 years, when only traditional risk factors were included in the models. The addition of imaging variables helped maintain the prediction ability beyond middle age.

### Multimodal Imaging Markers for Dynamic Prediction

This work is unique as it is among the first that incorporates high-dimensional longitudinal imaging markers from multiple modalities collected with high levels of sparsity (a high percentage of missing values) and irregularity (non-uniform time intervals between measurements) for dynamic prediction of CVRD. Many previous studies have limited the use of imaging data in prediction models, using only cross-sectional data or a few variables, or only including complete data. Simple imputation methods are often used to deal with sparsity and irregularity (mean/median imputation or last observation carried forward),^23^ but these can introduce bias and do not fully capture the information in longitudinal data. In this work, we employed Dynamic-DeepHit which was capable of dealing with data of high sparsity and irregularity,^11^ and thus could overcome the aforementioned challenges and better capture the rich information to improve risk estimation.

We showed that the inclusion of longitudinal multimodal imaging markers led to 0.03-0.05 increase in C-index and iAUC compared to not using imaging markers. It is worth noting that the imaging data collected in CARDIA was highly sparse and irregular (only available in two or three follow-up exams, collected in a small subset of participants (Table S1). The various missingness patterns reflected the nature of real-world data as not all information from a past patient visit will be collected in the current visit. We argue that, despite the variable missingness rates, longitudinal multimodal imaging markers still improved prediction up to 5%. More complete and frequently collected data will likely yield greater improvement in risk estimation.

This study found that the predictive accuracy of a model for cardiovascular disease risk dropped after the mean age of the participants reached 45 years old, especially when using only traditional risk factors. The inclusion of multimodal imaging markers helped stabilize the predictive accuracy and prevented a decline of 6-7% over 13 years. The decline in predictive performance when using only traditional risk factors may be due to several factors. First, traditional risk factors may be less effective for predicting 10-year cardiovascular risk in people entering midlife despite their demonstrated usage for longer-term prediction. Previous studies from our group have shown that traditional risk factors were not among the top predictors for short-term prediction in an older population (MESA cohort, mean age = 62),^7^ and also in the CARDIA population.^24^ Second, some non-traditional risk factors such as mental health, alcohol abuse, and other lifestyle factors, were not included in our prediction models. Studies have shown that cumulative effects of stress and alcohol contributed to worsening cardiovascular health.^25–27^ Third, health tends to decline starting in middle-age, when many changes occur in the body, making it more challenging to predict cardiovascular disease risk at this age. For example, at this age range, menopause often begins and the aortic root could enlarge and dilate, which have been shown to negatively affect cardiovascular functions and metabolism.^28,28^ More generally, metabolic syndrome in those 40–59 years of age were about three times as likely to happen as in those 20–39 years old.^29^

In this regard, the decline in predictive performance in using only traditional factors further highlighted the role of multimodal imaging markers. Even though the traditional risk factors are fundamental to the genesis and progression of CVRD, multimodal imaging markers can pick up physiologic signals that are closer to disease initiation and closer to adverse outcomes. Furthermore, imaging markers can capture signals from some of the cumulative effects of insults to the body that were not captured by traditional risk factors. For example, coronary calcification from CAC/CT has demonstrated the proatherogenic effects of heavy alcohol consumption since young adulthood.^30^ CAC/CT variables consistently ranked in the top predictors of outcome in our models (Table 2). Signals signifying the changes in the body at middle age could also be recognized by longitudinal imaging, for example, aortic root enlargement captured by Echo and was among the top-6 predictors of outcome at year 33, when the average participants’ age was 58. In addition, the importance of age decreased over time while the importance of the imaging markers increased (Table 2), suggesting that vascular age captured by imaging may be more relevant than chronologic age. Overall, the included multimodal longitudinal imaging markers stabilized the decrease in prediction accuracy but may not have captured all relevant information. Adding more diverse, high-quality multimodal data may be necessary to further improve prediction in this age group.

### Importance of imaging subsets

In this work, we quantified the importance of imaging markers as whole variable subsets/imaging modalities in addition to looking at variable-level importance. We also assessed spatial importance (in one exam) and temporal importance (across exams). We found that CT and Echo variables were consistently among the most important predictors. Specifically, within Echo variables, left ventricular dimensions, ventricular septal thickness, aortic root measurements, and circumferential peak strain were among the most important. These variables are also reportedly among the top predictors in other large-scale studies.^31^ For CT variables, markers from coronary artery calcium (CAC) scans were consistently among the top predictors, adding to the growing evidence in the literature about the importance of CAC. Abdominal aortic calcium variables such as the number and size of lesions of the abdominal aorta and common iliac aorta are also among the top 30- 50 predictors. Additionally, intermuscular adipose tissue (IMAT) measured by CT also consistently presented in the top 30, agreeing with previous reports showing IMAT associated with increased subclinical atherosclerosis independent of traditional cardiovascular disease risk factors and other adipose depots.^5^

In the early years, Echo markers, specifically markers of hypertension (such as septal thickness, LV volume and dimension), contributed the most to outcome prediction. However, in later years, CT markers played a larger role in prediction (Figures 3, 4, and Table 2). This suggests that at a young age, hypertension is the main driver of CVD, whereas at middle age, markers of atherosclerosis become the main driver, which can be more efficiently captured by CT/calcium scoring. Variables from the other subsets (e.g., DEXA, CARTD) contributed weakly to the prediction. Regarding brain MRI markers, total brain volume, including gray matter, white matter and cerebral spinal fluid and abnormal tissue volumes, primarily in white matter were among the top 15-20, and overall brain markers helped improve CVRD prediction in the immediate years after they were added. Previous studies have reported that cardiovascular risk burden is associated with cognitive decline, structural brain differences, and brain age.^32–35^ However, most studies show that CVD risk factors predict or are associated with brain structure and function,^33,35,36^ and not the other direction. Therefore, the brain MRI measures may reflect already accumulated CVD risk factors and therefore provide extra information on the severity of the risk factors further improving CVRD prediction.

### Algorithmic consideration

In our study, we compared several dynamic survival analysis algorithms to identify the best technique to handle sparse and irregular imaging data. Among the techniques tested, machine learning methods were superior to the Extended Cox model. The best-performing models were those using Dynamic-DeepHit trained on unimputed data, which can directly handle sparse, high dimensional, and irregular data and provide true dynamic prediction. LTRCforest trained on imputed data performed on par with Dynamic-DeepHit but was not a true dynamic prediction algorithm and required imputation and more computational time. Therefore, Dynamic-DeepHit may be the most suitable algorithm for dynamic prediction.

### Limitations

Our study has several limitations. The data collection started in 1985 in a biracial population and followed through for 30 years describing a certain cohort experience. Caution must be exercised when generalizing to other races and to the current population, as there may have been shifts in population characteristics over time. Second, external validation is challenging because long-term follow-up studies of young adults with extensive phenotyping like CARDIA are sparse.

Third, as noted, many imaging markers in CARDIA are highly sparse and irregularly collected, whereas quantification of longitudinal multimodal imaging utility would improve with complete data. Despite that, the inclusion of sparse and irregular multimodal imaging data still significantly improved prediction. Finally, the collection of repeated multi-modal imaging is practically possible mainly in well-resourced health facilities.

## Conclusions

We show that longitudinal multimodal imaging data readily collected from follow-ups can improve CVRD dynamic prediction. Echocardiography measured early can capture hypertension status and provide a good prediction for long-term risk estimation, while CT/calcium scoring variables carry atherosclerotic signatures that benefit more immediate risk assessment starting in middle-age.

## Data Availability

The CARDIA dataset can be requested via the study website https://www.cardia.dopm.uab.edu/. CARDIA study data are available to affiliated and non-affiliated investigators. See the study website for further details: http://www.cardia.dopm.uab.edu/invitation-to-new-investigators.

## Acknowledgments

We thank the CARDIA committee for reviewing the scientific content of this manuscript. The views expressed in this manuscript are those of the authors and do not necessarily represent the views of the National Heart, Lung, and Blood Institute; the National Institutes of Health; or the U.S. Department of Health and Human Services.

